# Assessing the Correlation of At-Home Audio Testing and In-office Uroflowmetry: Moving Towards a New Gold-Standard

**DOI:** 10.1101/2025.09.10.25335538

**Authors:** Raeven S. Grant, Kymora B. Scotland, Karan N. Thaker, Camille Watson, Yash Motwani, Myung-Shin Sim, Abigail Lavold, David F. Yao, Stephanie Pannell

**Affiliations:** Department of Urology, David Geffen School of Medicine, University of California-Los Angeles, Los Angeles, CA; Division of General Internal Medicine and Health Services Research, University of California, Los Angeles, CA

**Keywords:** Benign prostatic hyperplasia, Lower urinary tract symptoms, Uroflowmetry

## Abstract

**Objectives:** The incidence of lower urinary tract symptoms (LUTS) increases with age. Uroflowmetry, a diagnostic tool for LUTS, requires that patients travel to their urologist’s office. As audio-based uroflowmetry is being developed in the field, research is needed to evaluate whether it is equivalent to the gold standard, in-clinic uroflowmetry.

**Methods:** Men over age 18 with a chief complaint of LUTS conducted a standard in-office uroflowmetry and used the Emano Flow phone application for one week to conduct audio-based uroflowmetry. Pearson correlation coefficient (PCC) was utilized to assess the relationship between conventional and the mean of audio-based Qmax after adjusting for the effect of void volume by performing residualization.

**Results:** Thirty-two participants were recruited. On average, a patient’s home volume was 2.18x (SD 1.67) higher than their clinic volume. Prior to adjusting for void volume, conventional and the mean of multiple at-home maximum flow rates were not correlated (PCC=0.21, p=0.31). After adjusting for void volume, conventional and mean of multiple at-home Qmax were significantly correlated (PCC=0.41, p = 0.04).

**Conclusion:** Prior to adjusting for void volume, conventional and at-home maximum flow rates were not correlated. This can be explained, in part, by patients having one uncharacteristically low in-clinic on-demand void. Once we adjusted for void volume, office and at-home Qmax were correlated. As office Qmax may not accurately capture a patient’s natural voiding patterns, our findings underscore the potential value of at-home testing. Further work is needed to improve the tools that we use for diagnosing and evaluating LUTS.

## Introduction

Lower urinary tract symptoms (LUTS) are a grouping of symptoms related to the storage and emptying of urine, such as slow stream, need to strain, intermittency, and irritative symptoms such as urgency, frequency, nocturia, and dysuria^1^. The global burden of LUTS is quite high, with symptoms being experienced by an estimated 2.3 billion people^2^. Further, the incidence of LUTS increases with age. One study with 5000 male participants at least 65 years of age with no history of prostate cancer found that 46% of them experienced moderate to severe LUTS^3^. Patients with LUTS rate their health quality as poorer and report a marked impairment in activities of daily living. LUTS are thought to be due to a structural and/or functional abnormality in the bladder, bladder neck, prostate, sphincter, and/or urethra^4^. They are sometimes suggestive of benign prostatic hyperplasia (BPH) and subsequent bladder outlet obstruction, as well as several other etiologies^5^.

Uroflowmetry is a simple, noninvasive diagnostic tool for LUTS^4^. Patients urinate into a device that measures the flow rate and volume per void. Uroflowmetry requires that patients travel to their urologist’s office and void on demand, sometimes without an adequate urge. This one-time measurement may not be characteristic of a patient’s typical voids for various reasons. Though currently considered the gold-standard, new technology is needed to capture data from a patients’ typical voids, as well as increase accessibility and empower patients.

Indeed, new technology is constantly being developed in the field of urology to optimize health outcomes. A 2020 study found that there were over 500 applications on the Apple App Store and Google Play Store ^6^. One type of application that can be found is audio-based uroflowmetry. Audio-based uroflowmetry uses machine learning technology to estimate urinary flow rates and generate similar curves to conventional uroflowmetry at low-cost^7–10^. In an internal validation study, the Emano Flow technology, used in this study, has already been shown to be accurate in the home setting. In the same void, the app was used simultaneously with conventional uroflowmetry. The absolute error in predicted instantaneous flow was only 3.07ml/s. This technology can provide clinicians more insight into patient’s voiding patterns in a more natural setting. Despite the promise of this technology, there is little research validating its clinical equivalence to conventional uroflowmetry.

## Methods

### Study Population

Adult males over the age of 18 years old were recruited from two urology clinics. Only males were included, as patients were required to stand to urinate while using the phone application. Patients were invited to participate in the study if they had a chief complaint of LUTS and a smartphone. There were no other urologic history-related inclusion or exclusion criteria. All participants provided written informed consent to the use of their in-clinic uroflowmetry and audio uroflowmetry prior to participation in the study. The study was approved by the University of California Los Angeles Institutional Review Board (23-1068-CR-001).

### Data Collection

Conventional uroflowmetry was performed in the clinic. Audio-based uroflowmetry was conducted using the Emano Flow software. Emano Flow uses a patented deep machine learning technology to measure urinary flow rate and generate a flow curve using sound as urine makes contact with the water in a toilet bowl. Patients downloaded the Emano Flow application on their smartphone. To facilitate analysis of multiple recordings, participants were instructed to record as many voids as they could for 7 days. The software processes this voiding data in real-time, allowing the data to be available immediately for researchers.

Participants also completed questionnaires on urologic medical and surgical history, as well as the International Prostate Symptom Score (IPSS).

### Data Analysis

Medians and interquartile ranges (IQRs) are reported for continuous variables. The primary outcome was the average maximum flow rate (Qmax) per void. To assess the relationship between home and office void performance, we calculated the Pearson correlation coefficient (PCC) between the mean Qmax from audio-based uroflowmetry (home void) and the Qmax from conventional in-clinic uroflowmetry. Because Qmax is known to depend on voided volume^11^, and because average voided volume in the clinic was much lower than at home, we applied a residualization approach to remove the effect of voided volume on Qmax. Specifically, we conducted separate linear regression models for office and home settings, modeling Qmax as the dependent variable and voided volume as the independent variable. For home voids, we used each participant’s mean Qmax and mean voided volume across all eligible recordings. The resulting residuals represent volume-adjusted Qmax values. We then computed the PCC between the residuals from the office and home regressions to evaluate the correlation of Qmax between settings after adjusting for voided volume. Both regressions were linear, and no extreme outliers were identified; therefore, all data points were retained in the analysis.

In addition to evaluating Pearson correlation, we conducted a Bland-Altman analysis to assess individual-level agreement between office and home Qmax values after adjusting for voided volume. Specifically, we used the residuals from separate linear regressions of Qmax on voided volume (performed independently for home and office data) to obtain volume-adjusted Qmax values.

We also considered an alternative analytic approach using mixed-effects models to account for repeated home voids per participant. However, in this framework, the relationship between Qmax and voided volume in home setting was nonlinear, prompting the use of spline-based residualization. Despite this flexible modeling strategy, we did not observe a statistically significant correlation between home and office Qmax residuals using this model.

## Results

### Participants

Thirty-two male participants were recruited as part of the study (**Table 1**). The median age of participants was 64.0 years (IQR 54.5 - 71.5). A total of 1,232 voids were recorded using the Emano Flow phone application. The number of home voids per patient ranged from 2 to 159 (median = 34.0 [21.0, 52.5]).

**Table 1:**
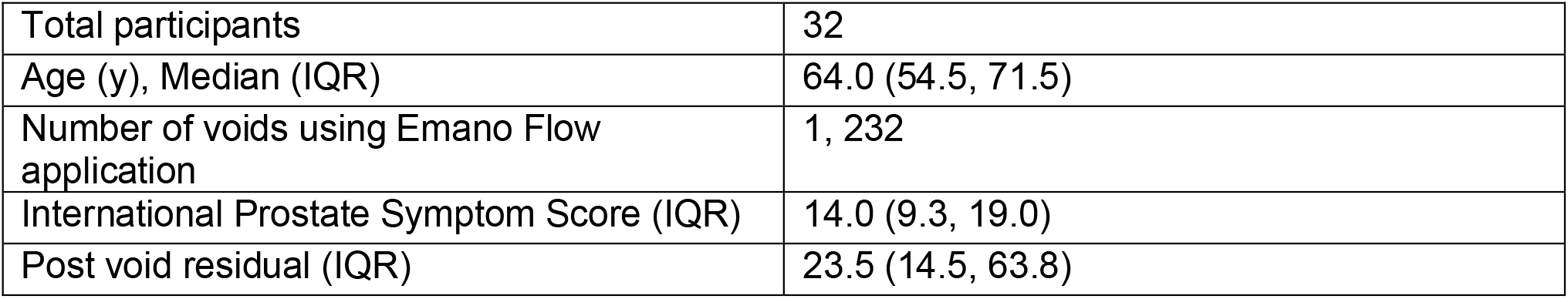
Patient Characteristics. Abbreviations: *IQR* interquartile range

### Correlation between Conventional Office-based Uroflowmetry and Audio-based Uroflowmetry

There was a linear relationship between office Qmax and voided volume (**Figure 1**). Audio-based uroflowmetry had a nonlinear relationship between Qmax and voided volume (**Figure 2**). The mean Qmax (mL/s) was 11.57 (SD= 7.38) and 17.05 (SD= 5.42) in-clinic and at-home, respectively. The mean voided volume (mL) was 168 (SD=146.69) and 238.68 (SD =98.54) in-clinic and at-home, respectfully. Initially, we found no significant correlation between in-clinic and mean of at-home Qmax (PCC=0.21; P = 0.31).

**Figure 1.**
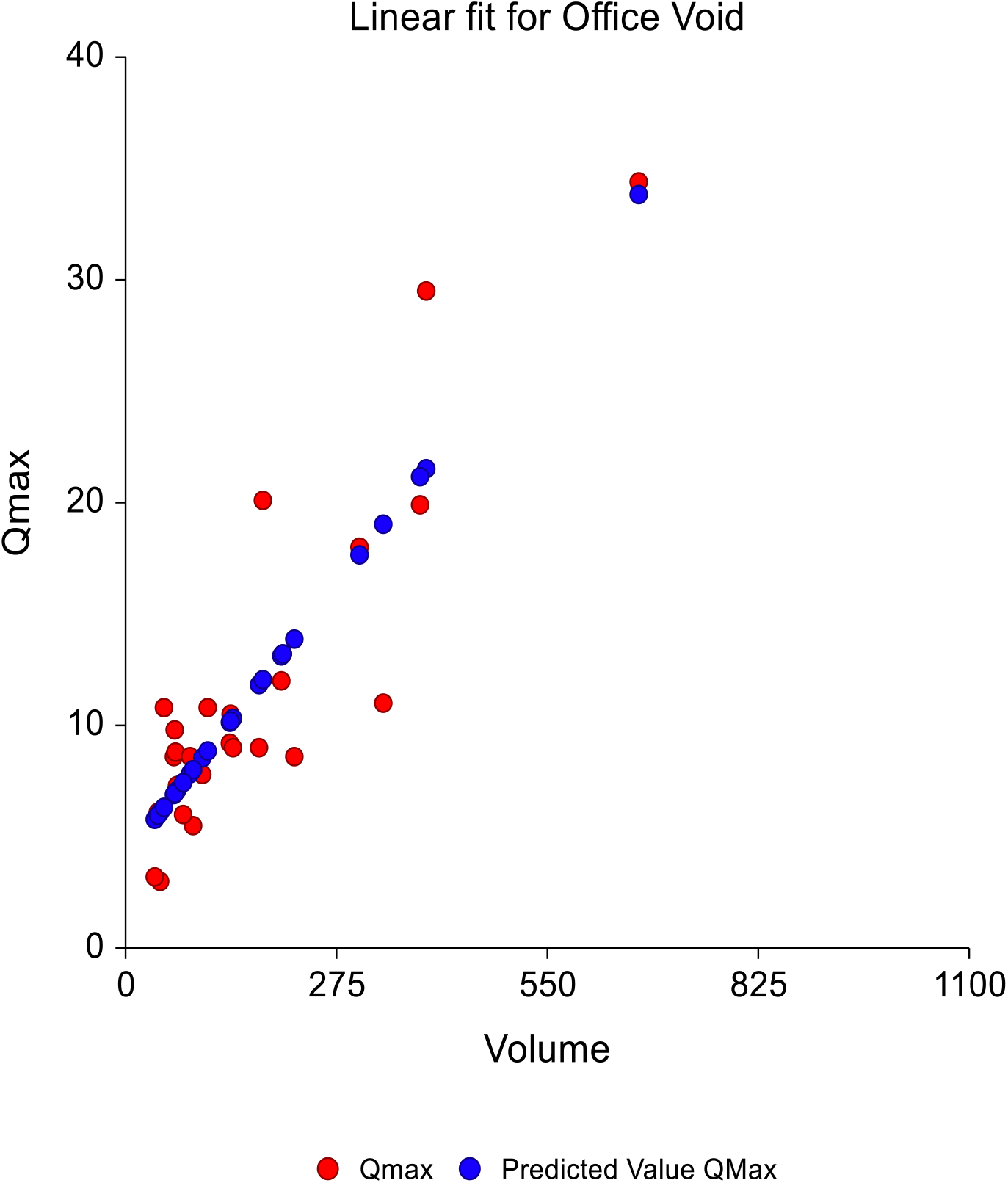
Fit showing linear relationship between Office Qmax and Volume

**Figure 2.**
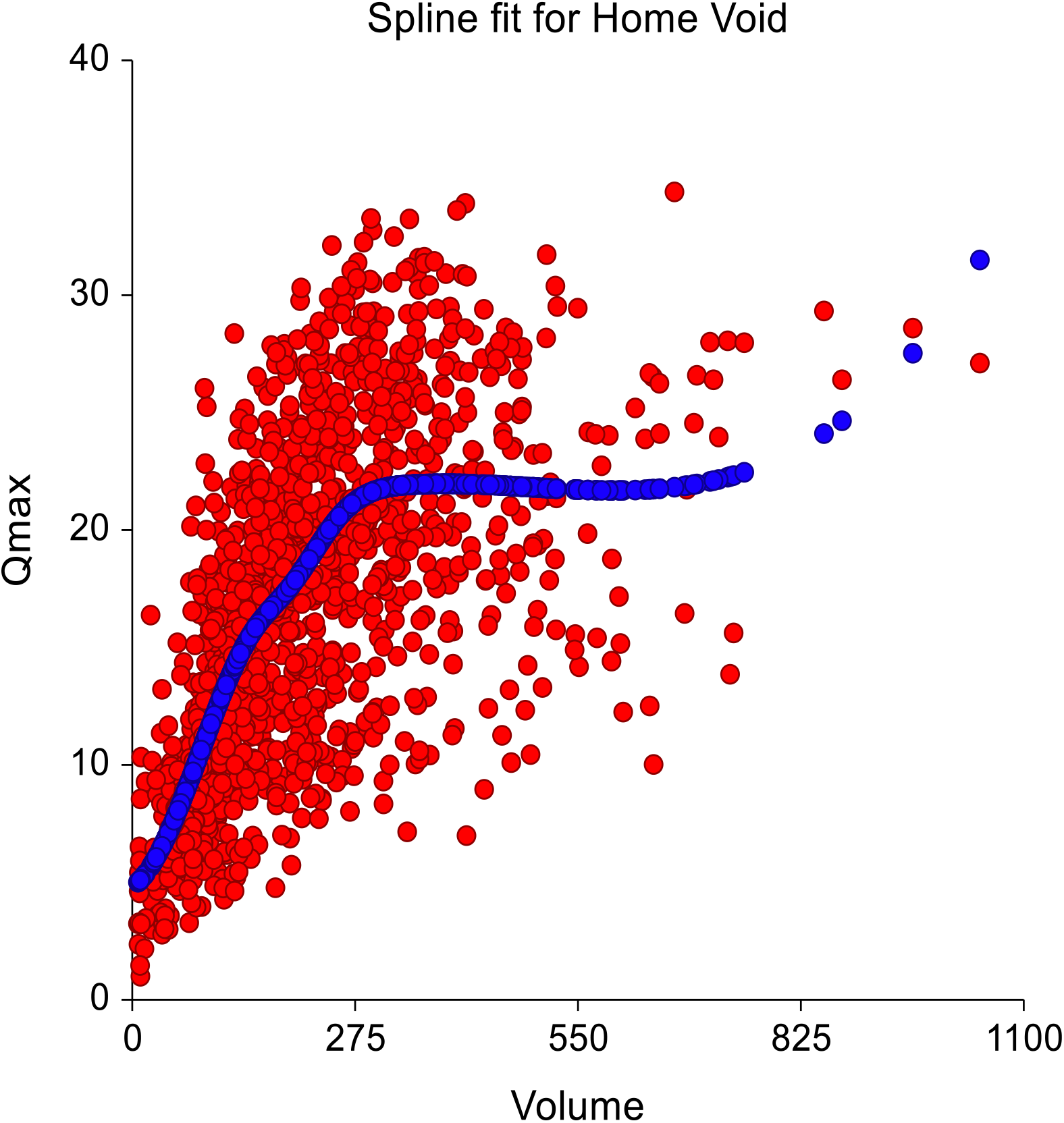
Spline fit showing nonlinear relationship between Home Qmax and Volume

The Bland-Altman plot showed a mean difference of −0.12 mL/s between office and home residuals, indicating no significant systematic bias (**Figure 3**). However, the 95% limits of agreement ranged from −8.02 to 7.78 mL/s, suggesting substantial variability in individual-level concordance between settings despite adjustment for voided volume.

**Figure 3.**
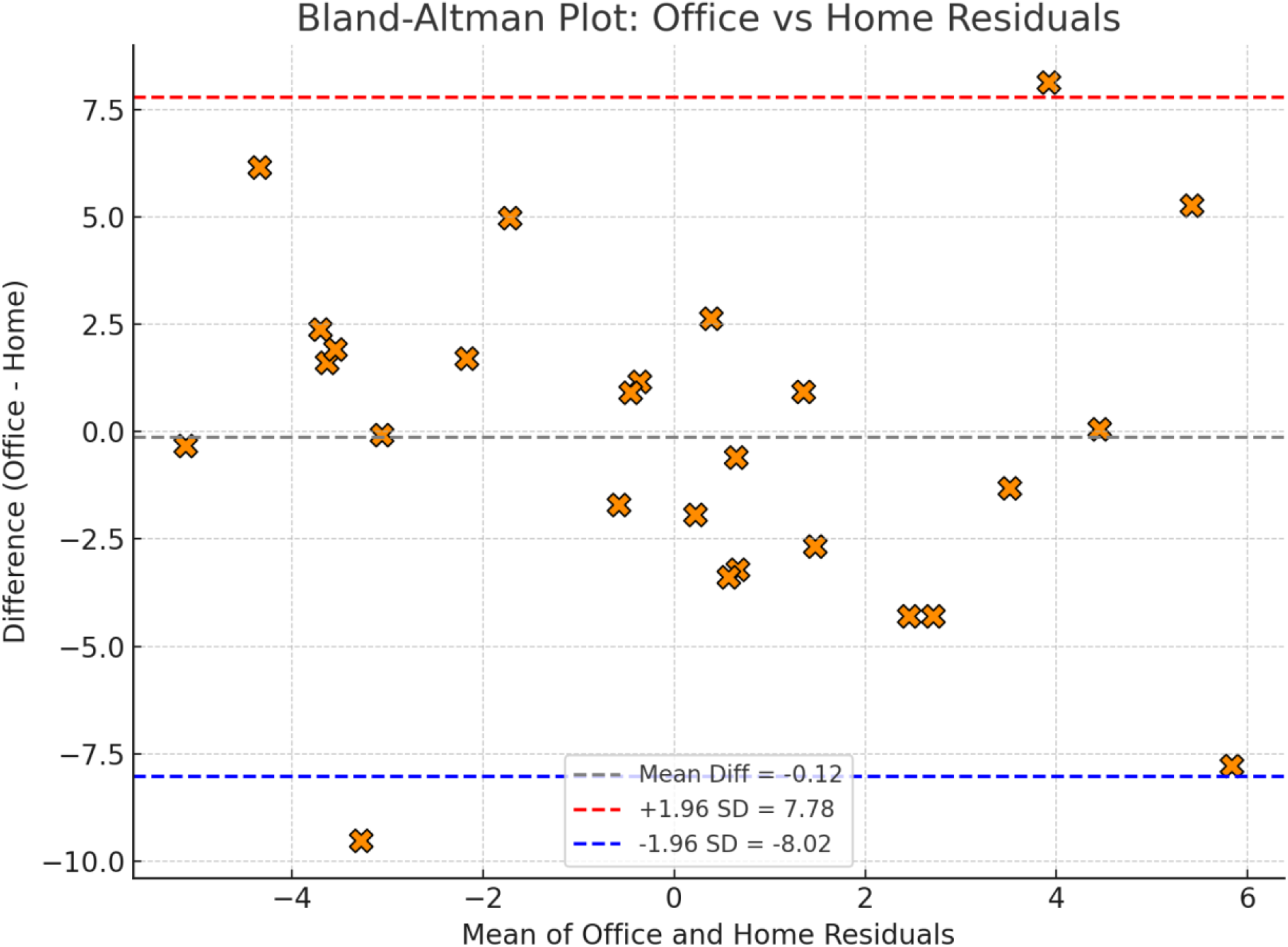
Bland-Altman Plot of Office and Home Residuals

We then computed residuals from the regression models and found a linear relationship (**Figure 4**). After performing residualization, we found a significant linear correlation between conventional and app-based uroflowmetry (PCC=0.41; 0.036).

**Figure 4:**
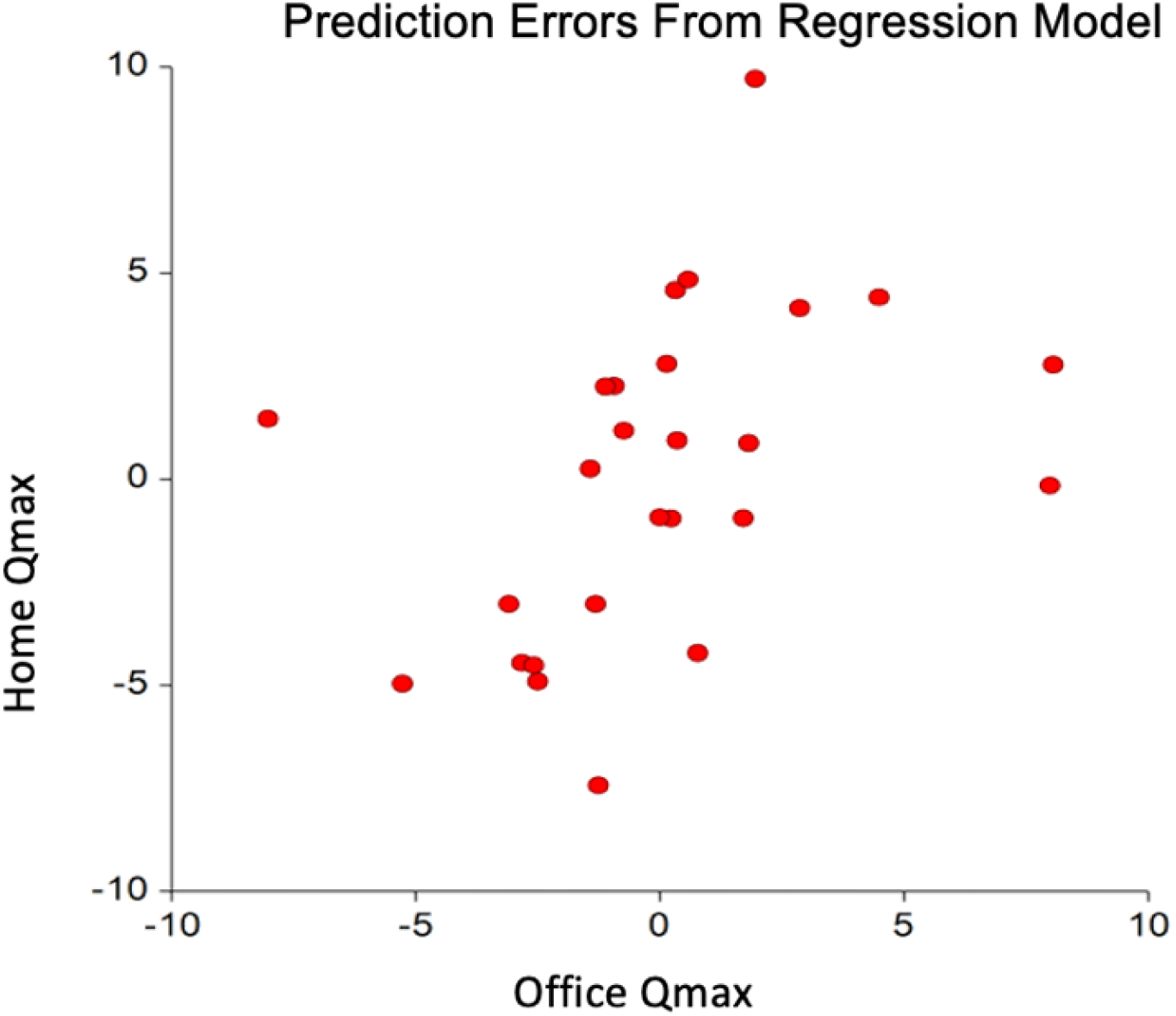
Linear relationship between home and office Qmax after adjusting for volume

## Discussion

Currently, in-clinic uroflowmetry is the gold standard for evaluating and diagnosing LUTS. New technology, including smartphone applications like Emano Flow, is being developed to optimize outcomes for patients. We sought to determine if data generated from the Emano Flow app are equivalent to that of the gold standard, in-clinic uroflowmetry. Initially we found no significant correlation between conventional and at-home Qmax. Once we removed the effect of voided volume, we found a significant correlation between the two. The results suggest that audio-based uroflowmetry can provide insight into patients voiding patterns at home, adding valuable data to in-office uroflowmetry.

There have been several studies exploring the in-clinic gold standard to audio-based uroflowmetry with mixed results. A study by Bladt and colleagues compared audio uroflowmetry, using similar technology to Emano Flow, to conventional uroflowmetry^12^. Interestingly, they found that 48% of patients had a Qmax in-clinic that fell out of the range of their at-home Qmax. As Qmax is dependent on voided volume^11^, the results in our study can also be in part explained by patients having an uncharacteristically low volume in-clinic void. Indeed, we found that at home voided volumes were over two times higher than the in-clinic volumes. A recent study by Dawideck and colleagues found that audio-based uroflowmetry was inconsistent with in-clinic uroflowmetry^7^. They concluded that this is due to variability in and difficulty standardizing sound signals. As the Emano Flow app has been validated, we believe this did not play a role in the results of our study.

Although in-clinic uroflowmetry has long been regarded as the gold standard for evaluating LUTS, the results of our study add to the literature discussing its limitations. Studies have shown that while in-clinic uroflowmetry is non-invasive and cost-effective, it creates an artificial environment in an unnatural setting^13^. Patients without the urge to void in clinic have to either wait, or void on demand, which deviates from their normal voiding patterns. Further, research has shown variability in urometric data collected throughout the day^14,15^. Thus, a one-time data point in the morning, for instance, might not be representative of patients’ voiding patterns throughout the day.

One limitation of our study is the exclusion of female participants. Currently the technology is only validated for the sound of urine hitting the liquid medium (toilet bowl) from a certain distance. It is possible that the results of the study would be different with the inclusion of those who sit to void. Once the technology has been further developed, a subsequent study should be done to evaluate the equivalence of the data generated by a cohort including females. Another limitation includes the inclusion of voids less than 150ml. Some studies suggest that these voids should be excluded, though we have included them in this study as they highlight the differences in voids produced at home versus in-clinic. Though our sample size is relatively small, a major strength of the study is the ability to study multiple, real-time voids over a week.

## Conclusions

Audio-based uroflowmetry can provide important insight into a patient’s natural voiding patterns, as it allows providers to review multiple voids, throughout the day, over a period of time. As LUTS negatively impact patients’ quality of life, access to real-time, real-life data is important for optimizing health outcomes for patients. Future work should be directed at improving the tools, both at-home and in-clinic, that are used to diagnose and evaluate LUTS.

## Data Availability

All data produced in the present study are available upon reasonable request to the author.

## Acknowledgements

We would like to acknowledge the Clinical and Translational Science Institute at UCLA for assistance with statistical analysis, and the Winston Research Grants Fund for financial support.

